# Comparison of Tooth Wear Among Young Sports Persons Involved in Individual and Team Sports- A Cross-Sectional Study

**DOI:** 10.1101/2024.11.05.24316808

**Authors:** Rangasamy RamyaDurga, KenniyanKumar SriChinthu, Harikrishnan Prasad, Muthusamy Rajmohan, Prema Perumal, Thuckanickenpalayam Ragunathan Yoithapprabhunath, S. Shanmuganathan

## Abstract

**BACKGROUND:** Tooth wear has multifactorial etiology & defined as non-self-reparable loss of hard tissue in the absence of dental caries or trauma. One of the valid aetiologies for pathological tooth wear is stress. During sports activity, athletes tend to clench their teeth involuntarily which results in tooth wear, especially in young sportspersons. Hence, we designed a study to evaluate and compare tooth wear among young sportspersons involved in individual and team sports.

**METHODS:** The study population comprised 300 subjects and were divided into two groups, group 1 (150 individuals involved in individual sports activity) & group 2 (150 individuals involved in team sport activity) The Tooth wear index was assessed based on Smith & Knight scoring criteria (1984).

**RESULTS:** 88.6% of study participants showed tooth wear in individual sports & 84% in team sports. Tooth wear was highest in the mandibular & maxillary central incisor of both individual & team sports individuals.

**CONCLUSION:** We found that both individual and team sports players showed tooth wear, especially in the incisal surface of mandibular & maxillary incisors, which might reflect the underlying stress or anxiety. This study is the first of its kind to evaluate and compare tooth wear among young sportspersons involved in individual and team sports in India.

## INTRODUCTION

Tooth wear is a non-self-reparable, multifactorial condition and is defined as the loss of hard tissue despite dental caries or trauma. The prevalence of tooth wear is increasing globally due to psychological and lifestyle factors. Physiological tooth wear is normal and is thought to reflect the aging process in humans. When the rate of tooth wear exceeds what is normal for the patient’s age, then it is considered pathological wear. Pathological tooth wear will necessitate dental treatment due to the manifestation of its symptoms. Excessive tooth wear results in dentin exposure, leading to dentinal hypersensitivity followed by pulpal and periapical inflammation, eccentric occlusion, trauma from occlusion, temporomandibular joint disorder and subsequently resulting in reduced chewing function.[1] One of the valid aetiologies for pathological tooth wear is stress.

During sports activity, athletes tend to clench their teeth involuntarily, which results in tooth wear, especially in young sports persons. Athletes should maintain good dental health to perform to the best of their abilities because oral disorders can have a direct impact on their general health. [2,3] Sports dentistry is an emerging field of dentistry in developed countries that aims to avoid injuries and enhance sports performance by examining the effects of dental health on professional and amateur players’ performance. It mainly includes the prevention and management of sports-related orofacial and dental injuries. [4,5]

The field of sports in India has its well-known importance and it is home to a diverse population playing several sports.[6] However, special training on dental and oral health maintenance during sports activity has not been emphasized, and awareness about the same is lacking among sports individuals when compared to any other developed country. To date, no research has been done on the prevalence of tooth wear among young athletes in the Indian community. Therefore, we designed a study to evaluate and compare tooth wear among young sportspersons from the South Indian population involved in individual sports activity and team sports activity.

### METHODOLOGY

A total of 300 participants were included in the study. The sample size was calculated using G-power software version 3.1.9.2. Participants were categorized into two groups - Group 1 (Comprising 150 individuals involved in individual sports activity-Silambam, athletics, wrestling, squash, taekwondo, badminton, and cycling) and Group 2 (Comprising 150 individuals involved in team sports activity - football, kho-kho, volleyball, basketball, cricket, handball, hockey, and kabaddi).

Only healthy sports individuals studying or working in a physical education institute, or sports clubs located in and around our district were considered for inclusion. Irrespective of their gender, those aged between 18 and 25 years and involved in practicing sports for a minimum of 2 years were involved in the study. Individuals undertaking orthodontic correction or with the habit of bruxism, and individuals who frequently took carbonated drinks were excluded from the study.

The Institutional Review Board approved our study design. Institutional Ethical Committee clearance was obtained (IEC no. 333/KSRIDSR/IEC/2022) before the commencement of the study. The procedure & confidentiality of the study were described in their local vernacular and due consent was obtained from the participants before the commencement of the work. The tooth wear of the individual was evaluated by using the Tooth Wear Index. The assessment was based on scoring criteria laid out by Smith & Knight (1984). Tooth surfaces were dried using cotton and examined for dental wear. Four surfaces examined were, Incisal (I) or Occlusal (O), Cervical (C), Labial (L) or Buccal (B), Palatal(P), or Lingual (L). According to the criteria, score 0 represents no loss of enamel surfaces on O/I/B/L and no change in the contour of C, score 1 represents the loss of enamel surfaces on I/ O /B/L and the slight loss of contour of C, score 2 denotes enamel loss & exposing dentin for less than 1/3rd of the surfaces on O/I/B/L and defect less than 1mm deep on C, score 3 denotes loss of enamel exposing the dentin for more than 1/3rd of the surfaces on O/I/B/L and defect 1 to 2mm deep on C and score 4 denotes complete loss of enamel, dentin on O/I/B/L and pulp exposure and defect more than 2mm deep on the contour of C. Restored & carious tooth surfaces were excluded. The obtained scoring was recorded in the data sheet for statistical analysis.

## RESULTS

Out of 300 study participants, 243 (81%) were males & 57 (19%) were females. 88.6% of study participants showed tooth wear in individual sports & 84% in team sports. [Table-1]

**Table 1.** Distribution of tooth wear.

| **Tooth wear** | **Individual Sport**<br>**(n=150)** | **Team Sport (n=150)** |
| --- | --- | --- |
| Yes | 133 (88.6%) | 126 (84%) |
| No | 17 (11.3%) | 24 (16) |

The frequency of tooth wear (Score 1) among individual sports persons was highest in the incisal surface of the maxillary central incisor followed by the mandibular central & lateral incisor. Score 2 was noticed higher in the incisal surface of the mandibular central incisor followed by the mandibular lateral incisor & maxillary central incisor. Score 3 was noticed only in the incisal surface of the mandibular canine. [Table-2]

**Table 2:**
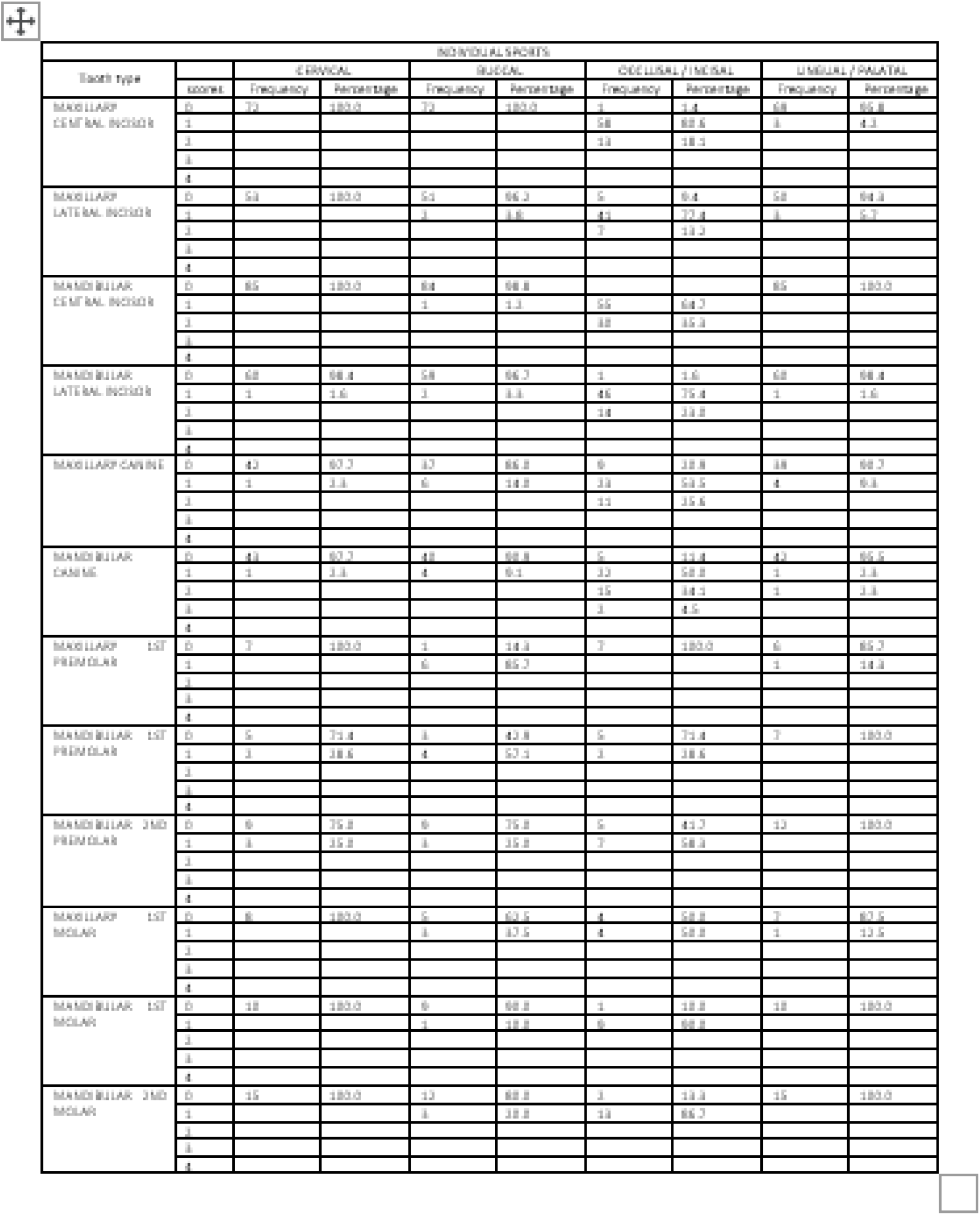
Frequency of tooth wear distribution in individual sports persons.

The frequency of tooth wear (Score 1) among team sports persons was highest in the incisal surface of the mandibular central incisor followed by the maxillary central & mandibular lateral incisor respectively. Score 2 was noticed higher in the incisal surface of the mandibular central incisor followed by the maxillary central & lateral incisor. Score 3 was noticed only in the incisal surface of the maxillary central incisor. [Table-3]

**Table 3.** Frequency of tooth wear in team sports persons.

| TEAM SPORTS |  |  |  |  |  |  |  |  |  |
| --- | --- | --- | --- | --- | --- | --- | --- | --- | --- |
| Tooth type | score | CERVICAL |  | BUCCAL |  | OCCLUSAL / INCISAL |  | LINGUAL / PALATAL |  |
|  |  | Frequency | Percentage | Frequency | Percentage | Frequency | Percentage | Frequency | Percentage |
| MAXILLARY CENTRAL INCISOR | 0 | 48 | 98.4 | 58 | 100.0 | 3 | 3.0 | 45 | 94.3 |
|  | 1 | 1 | 1.6 |  |  | 61 | 88.6 | 4 | 5.8 |
|  | 2 |  |  |  |  | 5 | 7.2 |  |  |
|  | 3 |  |  |  |  | 1 | 1.6 |  |  |
|  | 4 |  |  |  |  |  |  |  |  |
| MAXILLARY LATERAL INCISOR | 0 | 58 | 100.0 | 58 | 100.0 | 3 | 5.2 | 56 | 94.7 |
|  | 1 |  |  |  |  | 61 | 81.6 | 1 | 5.8 |
|  | 2 |  |  |  |  | 5 | 13.2 |  |  |
|  | 3 |  |  |  |  |  |  |  |  |
|  | 4 |  |  |  |  |  |  |  |  |
| MANDIBULAR CENTRAL INCISOR | 0 | 89 | 95.8 | 58 | 100.0 |  |  | 88 | 100.0 |
|  | 1 | 6 | 6.3 |  |  | 64 | 85.7 |  |  |
|  | 2 |  |  |  |  | 14 | 18.0 |  |  |
|  | 3 |  |  |  |  |  |  |  |  |
|  | 4 |  |  |  |  |  |  |  |  |
| MANDIBULAR LATERAL INCISOR | 0 | 10 | 90.3 | 58 | 100.0 | 8 | 45.0 | 56 | 100.0 |
|  | 1 | 6 | 14.8 |  |  | 61 | 87.0 |  |  |
|  | 2 |  |  |  |  | 8 | 11.2 |  |  |
|  | 3 |  |  |  |  |  |  |  |  |
|  | 4 |  |  |  |  |  |  |  |  |
| MAXILLARY CANNINE | 0 | 37 | 97.4 | 58 | 100.0 | 1 | 3.0 | 56 | 100.0 |
|  | 1 | 1 | 1.6 |  |  | 64 | 88.5 |  |  |
|  | 2 |  |  |  |  | 8 | 7.0 |  |  |
|  | 3 |  |  |  |  |  |  |  |  |
|  | 4 |  |  |  |  |  |  |  |  |
| MANDIBULAR CANNINE | 0 | 56 | 94.3 | 58 | 100.0 | 3 | 15.0 | 56 | 100.0 |
|  | 1 | 1 | 5.8 |  |  | 64 | 88.5 |  |  |
|  | 2 |  |  |  |  | 2 | 5.0 |  |  |
|  | 3 |  |  |  |  |  |  |  |  |
|  | 4 |  |  |  |  |  |  |  |  |
| MAXILLARY PREMOLAR 1<sup>st</sup> | 0 | 1 | 100.0 | 1 | 100.0 |  |  | 1 | 100.0 |
|  | 1 |  |  |  |  | 1 | 100.0 |  |  |
|  | 2 |  |  |  |  |  |  |  |  |
|  | 3 |  |  |  |  |  |  |  |  |
|  | 4 |  |  |  |  |  |  |  |  |
| MAXILLARY PREMOLAR 2<sup>nd</sup> | 0 | 1 | 100.0 | 1 | 100.0 |  |  | 1 | 100.0 |
|  | 1 |  |  |  |  | 1 | 100.0 |  |  |
|  | 2 |  |  |  |  |  |  |  |  |
|  | 3 |  |  |  |  |  |  |  |  |
|  | 4 |  |  |  |  |  |  |  |  |
| MANDIBULAR PREMOLAR 1<sup>st</sup> | 0 | 4 | 100.0 | 4 | 25.0 | 1 | 15.0 | 4 | 100.0 |
|  | 1 |  |  | 1 | 25.0 | 4 | 15.0 |  |  |
|  | 2 |  |  |  |  |  |  |  |  |
|  | 3 |  |  |  |  |  |  |  |  |
|  | 4 |  |  |  |  |  |  |  |  |
| MANDIBULAR PREMOLAR 2<sup>nd</sup> | 0 | 6 | 100.0 | 1 | 87.5 | 1 | 15.0 | 7 | 87.5 |
|  | 1 |  |  | 1 | 12.5 | 4 | 15.0 | 1 | 12.5 |
|  | 2 |  |  |  |  |  |  |  |  |
|  | 3 |  |  |  |  |  |  |  |  |
|  | 4 |  |  |  |  |  |  |  |  |
| MAXILLARY 1<sup>st</sup> MOLAR | 0 | 1 | 100.0 | 1 | 100.0 |  |  | 1 | 100.0 |
|  | 1 |  |  |  |  |  |  |  |  |
|  | 2 |  |  |  |  | 1 | 100.0 |  |  |
|  | 3 |  |  |  |  |  |  |  |  |
|  | 4 |  |  |  |  |  |  |  |  |
| MANDIBULAR MOLAR 1<sup>st</sup> | 0 | 11 | 100.0 | 11 | 81.7 | 1 | 8.3 | 11 | 100.0 |
|  | 1 |  |  | 1 | 9.1 | 11 | 81.7 |  |  |
|  | 2 |  |  |  |  |  |  |  |  |
|  | 3 |  |  |  |  |  |  |  |  |
|  | 4 |  |  |  |  |  |  |  |  |
| MANDIBULAR MOLAR 2<sup>nd</sup> | 0 | 4 | 100.0 | 4 | 100.0 | 1 | 15.0 | 4 | 75.0 |
|  | 1 |  |  |  |  | 4 | 15.0 | 1 | 25.0 |
|  | 2 |  |  |  |  |  |  |  |  |
|  | 3 |  |  |  |  |  |  |  |  |
|  | 4 |  |  |  |  |  |  |  |  |

Since the severity of tooth wear was found to be highest in mandibular & maxillary central incisors of both individual & team sports individuals, a comparison of tooth wear among them was done by using Mann – Whitney U test. There was a statistically highly significant difference in the tooth wear of the mandibular central incisor between team & individual sports, especially in the incisal & cervical surfaces. [Table-4]

**Table 4.** Comparison of tooth wear in maxillary & mandibular central incisors between individual sports and team sports using Mann-Whitney U test.

| **Maxillary central Incisor surface** | **P-value Mann-Whitney U Test** | **Mandibular central Incisor surface** | **P-value Mann-Whitney U Test** |
| --- | --- | --- | --- |
| Cervical surface | .307 | Cervical surface | .004 |
| Labial surface | 1.000 | Labial surface | .283 |
| Incisal surface | .097 | Incisal surface | **.001** |
| Lingual/Palatal surface | .657 | Lingual /Palatal surface | 1.000 |

## DISCUSSION

Athletes’ success is based on several variables working together to achieve optimal performance. Failure could occur if any part of the training process is ignored. [7]

Dental health plays a vital role in sports performance, maintaining proper oral hygiene is important which could allow athletes to maintain their training and competition routines without hindrance in their sports plans due to dental issues. Therefore, it is highly suggested that athletes’ medical follow-ups should include an assessment of their dental status [7,8]

Oral health status and awareness among sports individuals have been surveyed in different regions of the world. Oral health plays a major role in the physical and emotional well-being of sports persons.[6] The most prevalent oral health problems in athletes are tooth clenching and bruxism or tooth grinding which results in tooth wear. A total of 300 participants were included in our study, of whom 150 were involved in individual sports activities and the remaining 150 in team sports activities. The age group selected was 18 to 25 years old to avoid the physiologic tooth wear that occurs because of the aging process.

In the present study, tooth wear was assessed by the tooth wear index proposed by Smith and Knight (1984). We found that participants involved in individual sports showed higher tooth wear (88.6%) when compared to the participants involved in team sports (84%). Similar findings were reported by De la Parte A et al in 2021, where they observed a higher percentage of tooth wear in individual sports when compared to team/group sports in the Spain population.[8]

In the present study, tooth wear was noticed predominantly in the anterior teeth of both individual and team sports persons. The frequency of tooth wear among individual sports was higher in the incisal surface of the maxillary central incisor followed by the mandibular central & lateral incisor respectively. Similarly, the frequency of tooth wear among team sports was found to be higher in the incisal surface of the mandibular central incisor followed by the maxillary central & mandibular lateral incisor respectively.

The severity of tooth wear was highest in the incisal surface of the mandibular & maxillary central incisor of both individual & team sports. Hence comparison among them was carried out by the Mann-Whitney U test. We found a statistically highly significant difference in the tooth wear of the mandibular central incisor between team & individual sports, especially in the incisal surface. Overall, the severity of tooth wear was found to be highest in the mandibular incisors of individual sportspersons when compared to team sportspersons.

In the original study by K Sun et al in 2017, they found that tooth wear was higher in the incisal surface of mandibular incisors followed by the maxillary incisor in a mixed-age group among the Chinese population.[9] Since there is no other study available to evaluate tooth wear in different types of teeth in athletes, the results of the present study cannot be compared directly.

The possible reason behind the increased tooth wear in athletes might be anxiety before a game or match. The anxiety can make them grind or clench their teeth, which might result in tooth wear. The grinding of teeth reflects the physical manifestation of stress or anxiety. [10,11] As competitive sports persons devote more time to training and strenuous exercise, their psychological and physical stress levels rise, potentially affecting their dental health and overall performance. [6]

## CONCLUSION

We found that participants involved in individual sports showed higher tooth wear when compared to participants involved in team sports. The frequency of tooth wear was highest in the incisal surface of mandibular & maxillary incisors in both individual and team sports players. The severity of tooth wear was noticed more in individual sports persons which might reflect the underlying stress or anxiety. This is the first study of its kind to evaluate and compare tooth wear among young sportspersons involved in individual and team sports in India. Awareness among the Indian sports community about maintaining good oral health is essential. A separate specialty in sports dentistry is the need of the hour. Identifying tooth wear at the earliest and planning the appropriate preventive measures for the same may be of great benefit to sports individuals in the Indian population.

## Data Availability

All data produced in the present study are available upon reasonable request to the authors

## Acknowledgment

Nil

